# Navigating HPV Vaccination Challenges in Low-Resource Settings: Practical Strategies for Estimating the Size of Out-of-School Girls

**DOI:** 10.64898/2026.04.28.26351102

**Authors:** Soim Park, Erica N. Rosser, Leanne Zhang, Megan D. Wysong, Arman Majidulla, Marina Terada, Pamela J. Surkan, Joseph G. Rosen, Rupali J. Limaye

**Author notes:** **E-mail addresses:**. **Corresponding author:**Soim Park, 615 N. Wolfe St. Baltimore, MD, 21205, USA.

## Abstract

**Background:** Achieving the 2030 target of 90% human papillomavirus (HPV) vaccination coverage among girls by age 15 requires effectively reaching out-of-school (OOS) girls, particularly in low- and middle-income countries (LMICs) where most vaccine delivery occurs in school settings. This study explored how diverse countries define OOS girls and strategies for estimating the size of this population.

**Methods:** Between May and September 2024, we conducted semi-structured in-depth interviews with 32 HPV vaccine program stakeholders across six African and Asian LMICs with established HPV vaccination programs: Cambodia, Cameroon, Kenya, Malawi, Mozambique, and Uganda. Using hybrid inductive-deductive thematic analyses, we examined how countries taxonomize OOS girls as well as current practices for enumerating or estimating population sizes.

**Results:** Countries commonly defined OOS girls as those not enrolled in schools under the Ministry of Education’s purview, but in practice, this definition varied across contexts. National-level data (e.g., census, school enrollment rates) and household-level approaches (e.g., headcounts) were commonly used to enumerate in-school and OOS girls. However, the distinction between in-school and OOS girls was often blurred during HPV vaccination implementation and evaluation, limiting the operational utility of these methods. To address these challenges, some countries adopted alternative approaches, including proxy-based population estimates using historical or approximate data and the prioritization or areas perceived to have a higher presence of OOS girls. These alternative approaches were perceived as pragmatic compromises between accuracy, feasibility, and resource constraints.

**Conclusion:** Accurate enumeration of OOS girls is important for HPV vaccination but may not be feasible in resource-limited settings. Pragmatic, context-specific estimation methods can offer a feasible way to identify and reach OOS girls more effectively with HPV vaccination services.

## 1. Introduction

Cervical cancer remains a key driver of health inequities, disproportionately affecting women in low- and middle-income countries (LMICs) where access to prevention, screening, and treatment services remains limited [1–3]. In 2022, an estimated 660,000 new cervical cancer cases and 350,000 deaths were reported globally, with more than 90% of this burden occurring in LMICs [3]. Nearly all cervical cancer cases are attributable to persistent infection with high-risk strains of human papillomavirus (HPV) and are preventable through timely vaccination [3]. In 2020, the World Health Organization (WHO) launched a global strategy to eliminate cervical cancer as a public health problem, including a 2030 target to vaccinate 90% of girls against HPV by the age 15 [4]. This strategy, combined with expanded access to screening and treatment, is expected to avert over 14 million cervical cancer deaths in LMICs by 2070 [4,5]. However, while 155 countries had introduced HPV vaccination into their national immunization programs by December 2025, significant gaps in coverage still persist [6].

One critical challenge in HPV vaccination programs is reaching out-of-school (OOS) girls through routine immunization delivery platforms. In LMICs, HPV vaccination is primarily delivered through a school-based approach [7]. Although cost-effective and logistically efficient, this method systematically misses OOS girls—those not enrolled in formal education systems registered with the Ministry of Education [8,9]. Approximately 40 million girls aged 6–11 and 30 million girls aged 12–15 were OOS in 2023 globally—a population which overlaps with age-eligible cohorts for HPV vaccination [10]. Although the global number of OOS girls declined between 2000 and 2023 (from 12 million to 7 million for girls aged 6–15), the number living in sub-Saharan Africa has remained relatively steady. Between 2000 and 2023, the region’s share of the global OOS children increased from 33% to 51% for children at primary level (aged 6–11), and from 26% to 50% for those at lower secondary level (aged 12–15) [10]. In contrast, southeastern Asia accounts for a relatively small and declining share of OOS girls, from 7% to 4% at primary level and from 9% to 6% at the lower secondary level over the same period [10]. For LMICs with a sizeable number of OOS girls, reaching these girls is imperative for HPV vaccine delivery.

Enumeration is essential for the effective implementation of vaccination programs. Without reliable enumeration, administrative coverage rates may be over- or under-estimated during evaluations, leading to potential gaps in immunization efforts [11]. Unlike other vaccine antigens, distinguishing between in-school and OOS girls is crucial for HPV vaccine planning and implementation in LMICs, given the predominance of school-based delivery [12]. To effectively reach OOS girls outside the school platform, a clear definition of this target population is the first step toward accurate enumeration. Administratively, the definition of OOS girls is standardized and refers to girls of official school age who are not enrolled in any formal education program under the Ministry of Education’s purview [10,13]. However, in practice, this definition often shifts during implementation. A recent review noted that inconsistent definitions and limited differentiation among OOS girls—those absent from school on vaccination day versus those not enrolled in school—complicate efforts to enumerate vaccine-eligible girls [12,14].

During the planning phase, accurately identifying target populations is essential for estimating and allocating resources based on actual needs [11]. Prior studies indicate that enumeration of target population remains challenging, even in high-income countries without universal personal identification numbers [15]. This challenge is more significant in resource-constrained settings, where the number of OOS girls in the community is often unknown [16,17].

Nevertheless, LMICs continue to enumerate and reach OOS girls with HPV vaccination services by developing and implementing strategies relevant to their contexts. Given the complex nature of vaccinating OOS girls, this study sought to explore how OOS girls are defined in HPV vaccination programs across six LMICs and to identify existing, feasible methods for quantifying this population.

## 2. Methods

### 2.1. Study Design

The study involved key informant interviews with stakeholders across five African and one Asian countries—Cambodia, Cameroon, Kenya, Malawi, Mozambique, and Uganda—selected to reflect variation in geographic region, HPV program maturity, and school enrollment contexts (see **Table 1**). Except for Cambodia, which introduced the HPV vaccine through a multi-age cohort (MAC) campaign in 2023, all other countries had at least three years of ongoing HPV vaccine programs at the time of the study. These countries had routinized their HPV vaccination, targeting 9- or 10-year-old girls, most of whom were enrolled in primary schools.

**Table 1.**
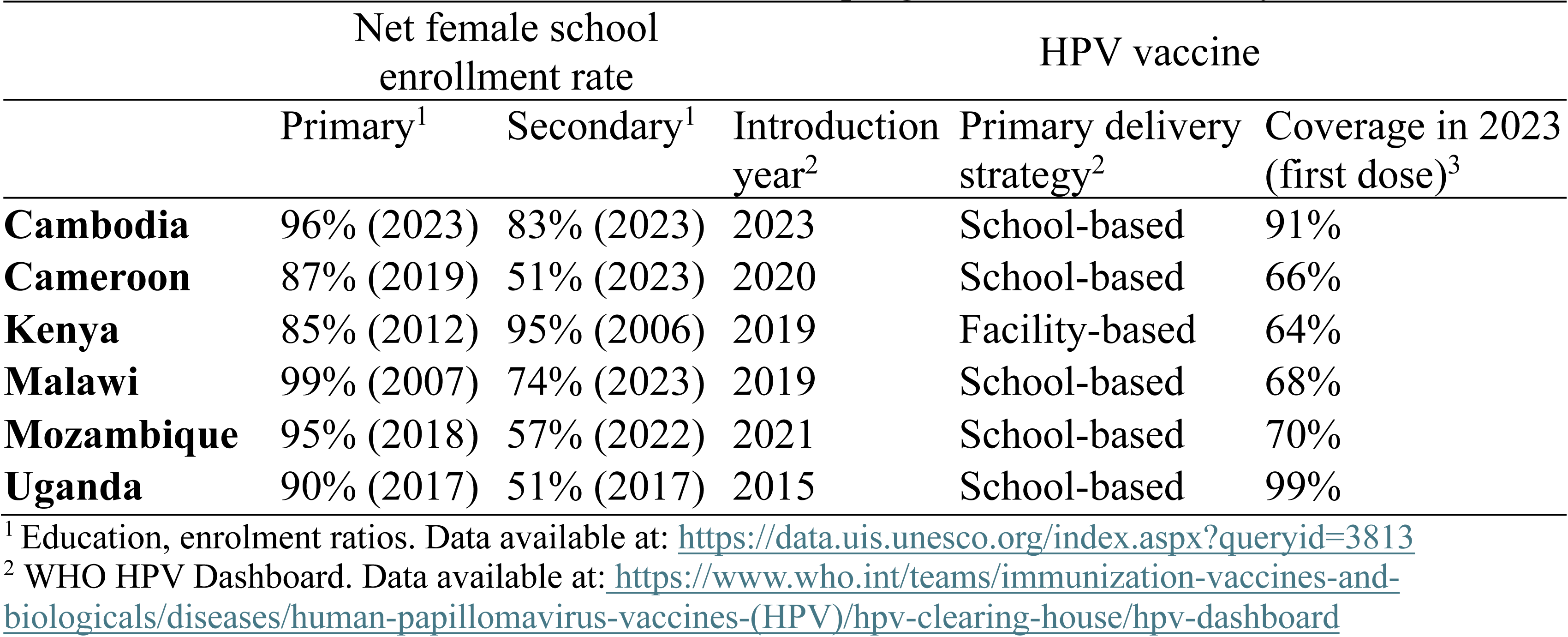
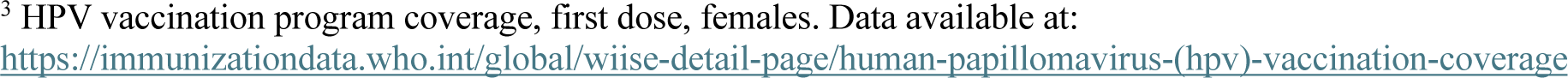
School enrollment and HPV vaccination program data across study countries.

A purposive sampling approach was used to recruit national and sub-national stakeholders involved in HPV vaccine policy, delivery, and implementation. These included representatives from Ministries of Health, Gavi-supported technical assistance partners, multilateral organizations, and civil society organizations. Eligible participants were aged 18 or older, had English- or French-language proficiency, and had professional affiliations with HPV immunization activities, particularly focusing on reaching OOS girls, in one of the study countries. Initial participants were identified through HAPPI Consortium technical assistance partner networks, with additional stakeholders recruited via snowball sampling based on referrals during interviews. In light of the richness of the interview data, we aimed to recruit 4-8 participants per country.

Out of 95 initially identified stakeholders, 82 were contacted by email, and 52 expressed interests in participating. Between May and September 2024, a total of 32 interviews were completed, including one paired interview at the request of the participants (**Figure 1**). The study protocol was reviewed by the Institutional Review Board at the Johns Hopkins Bloomberg School of Public Health and was deemed exempt from human subjects’ oversight.

**Figure 1.**
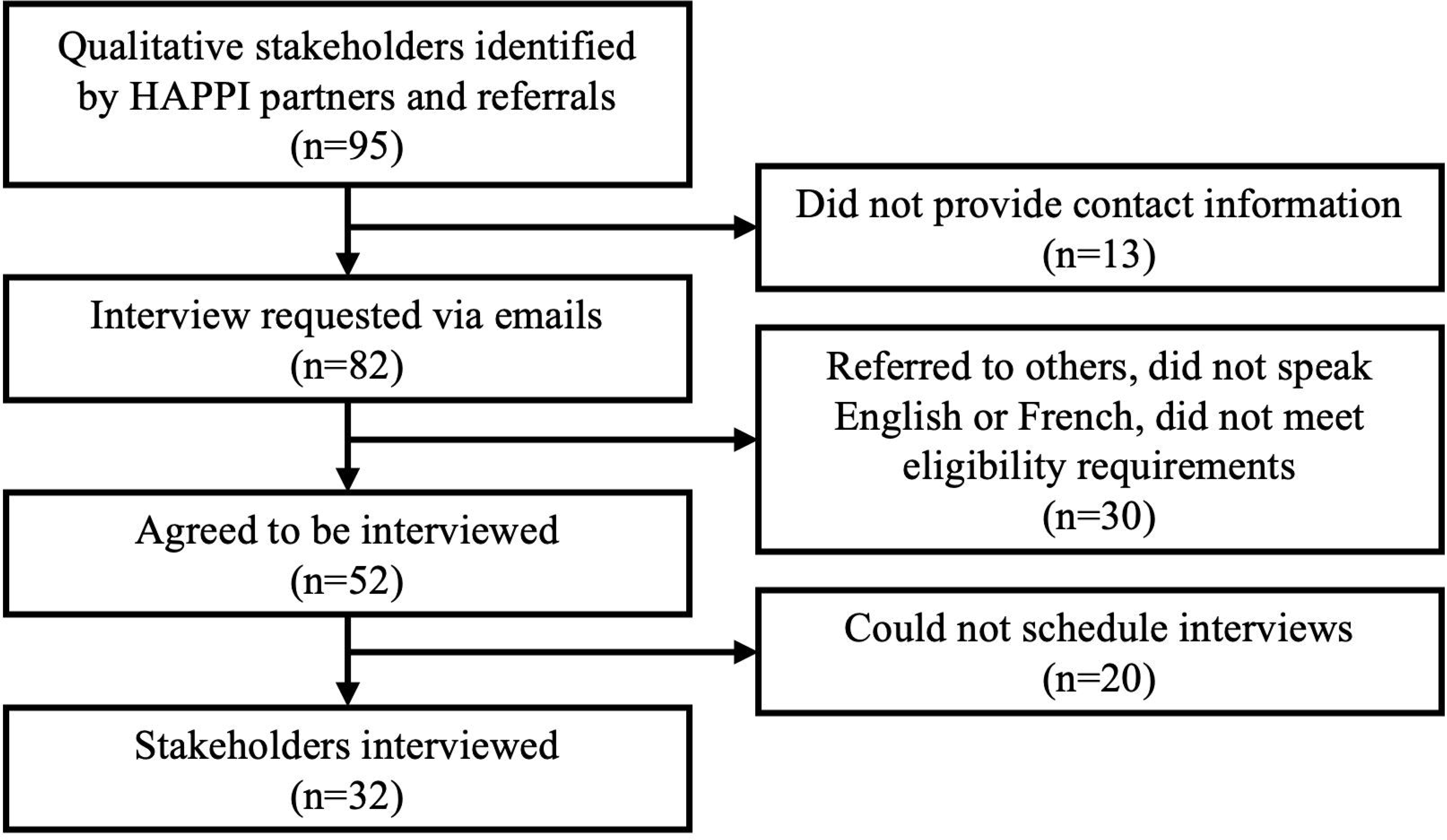
Participant recruitment flowchart.

### 2.2. Data Collection

Semi-structured in-depth interviews were conducted virtually between May and September 2024 by three researchers with graduate-level training in qualitative research methods. Participants were interviewed individually via Zoom audio-conferencing platform in their preferred language for 30 to 60 minutes. All interviews were audio-recorded after oral consent and professionally transcribed verbatim.

The interviews explored key dimensions of HPV vaccination efforts targeting OOS girls, including definitions and sub-populations of OOS girls, existing strategies to reach OOS girls with HPV vaccination services, and relevant implementation experiences including planning processes, adaptations, successes, and sustainability considerations. Participants were also probed on how immunization programs attempted to quantify or enumerate OOS girls and the role of local actors in these efforts. Interviewers were encouraged to probe for country-specific examples and to adapt the sequence or emphasis of questions based on the informant’s role and experience.

### 2.3. Data Analysis

A team-based thematic analysis approach was used to identify patterns across interviews and generate analytic categories related to the enumeration and vaccination of OOS girls. The research team developed a coding framework that combined inductive codes emerging from the data with deductive codes aligned with the study’s core domains, including implementation strategies, enumeration practices, contextual barriers, and stakeholder roles.

Coding was conducted iteratively by three team members, who regularly reviewed coded excerpts and refined code definitions through discussion. Coded data were then organized into a structured matrix to support within-case and across-case syntheses of emerging themes [18]. Horizontal analyses were conducted to compare findings across countries within each thematic area, while vertical analyses examined the interplay of multiple themes within each country. These steps supported the generation of locally understood definitions of OOS, their typologies, strategies for enumeration, and the recommendations presented in the subsequent sections. Atlas.ti Web (version 9.4.3) was used to facilitate data management, coding, and interpretation.

## 3. Results

We interviewed 32 stakeholders from six countries, ranging from 3 in Mozambique and 8 in Kenya. whom we interviewed included national immunization program officials, technical assistance partners, civil society representatives, and multilateral agency staff (**Table 2)**.

**Table 2.** Characteristics of interviewed HPV vaccination program stakeholders (*n* = 32).

| Country | Total | Affiliations |  |  |
| --- | --- | --- | --- | --- |
|  |  | MOH, EPI, and NITAG | TA partners and CSOs | Multilaterals |
| Cambodia | 5 | 0 | 3 | 2 |
| Cameroon | 8 | 4 | 4 | 0 |
| Kenya | 7 | 6 | 1 | 0 |
| Malawi | 4 | 0 | 3 | 1 |
| Mozambique | 3 | 2 | 1 | 0 |
| Uganda | 5 | 2 | 2 | 1 |

Abbreviations: MOH: Ministry of Health; EPI: Expanded Programme on Immunization; NITAG: National Immunization Technical Advisory Group; TA: Technical Assistance; CSOs: Civil Society Organizations.

### 3.1. Defining Out-of-School Girls

Across study countries, there was no single, universal definition of out-of-school girls. One stakeholder characterized OOS status as a binary distinction, “girls in school and girls out-of-school.” However, many stakeholders described more complex and contextually grounded understandings of who qualifies as OOS.

While in-school girls defined as those attending formal school, the presence of in formal, religious schooling systems in some countries complicated definition of OOS girls. Girls enrolled in madrasa or church-run institutions not recognized by national education systems were often omitted from official education records. In Kenya, stakeholders explained that students attending informal religious schools, which are not registered with the Ministry of Education, are technically considered OOS—even though they attend school daily. One Kenyan stakeholder noted, “Out-of-school [typically] means those who are not schooled at the moment. But we have different dynamics…[in communities] where the predominantly Muslim population is high. They are not in the formal school, [but] they are in Islamic schools.”

Others introduced the idea of temporality, differentiating between girls permanently excluded from schooling (e.g., due to school fees or social constraints) and those temporarily absent due to illness or social obligations. One stakeholder explained, “Out-of-school is a child who’s not able to go back to receive that education,” suggesting that extended school absence or educational attrition may be relevant to defining OOS status. In summary, for some, the definition of OOS girls is administratively rigid; for others, it is flexible and adopting to local educational, cultural, or economic contexts.

Many stakeholders described locally defined efforts to categorize OOS girls for programmatic purposes. Building on these heterogenous definitions, we defined a typology of OOS girls based on stakeholder accounts of who is most likely to be excluded from formal school enrollment and, thus, from school-based vaccination strategies. Four commonly reported drivers emerged: socioeconomic status, social norms, conflict and natural disasters, and disability.

- **Socioeconomic Exclusion:** Poverty and household financial constraints were cited as the most widespread drivers of school non-enrollment and attrition. Even where public education is nominally free, costs associated with transport, uniforms, or meals often complicate attendance and render school inaccessible. In some cases, girls were expected to support household income or provide domestic labor: *“[Girls] are dropping out because they need to support their families. Many families are farmers in rural areas, in rice fields or other farms, or [operate] personal shops or restaurants. And then many subpopulations [are] migrants, international or domestic migrant families, so they [girls] need to go with parents.”* – Cambodia
- **Social Norms:** Rigid gender norms governing girls’ roles in marriage, caregiving, and domestic labor were also linked to school dropout and non-enrollment. Some communities or households may prioritize preparing girls for early marriage, pregnancy, and domestic duties over pursuing their education, leading to increased rates of school attrition. Boys’ education may also be a higher priority than girls’ education, especially for families with limited or constrained financial resources. *“Culturally, women are to be at home, cooking, doing the house chores, caring for siblings, rather than going to school, because this is seen as men’s thing.”* – Mozambique Girls in specific religious communities may also attend informal religious schools that operate outside of the national education system.
- **Conflict and Natural Disasters:** Armed conflict and natural disasters were also cited as disrupting access to both education and healthcare services, particularly in Cameroon, Mozambique, and Uganda. Girls in internally displaced populations or refugee settlements faced additional structural barriers: *“With the war in the North-West and in the South-West, there are also more and more girls who cannot go to school because of the security context that prevails in these two regions.”* – Cameroon
- **Disability:** Girls with physical or cognitive disabilities were widely described as excluded from formal education due to infrastructural inaccessibility or insufficient support. *“When you look at the out-of-school girls, the most common is what we identify as the people enabled differently, the so-called disability. They are people with cerebral palsy. They are disabled. The parents, due to stigma, they are hiding them at home. So, these girls we are seeing out-of-school, but who are also not getting the vaccine.”* – Kenya

This typology reflects overlapping and intersectional vulnerabilities, with many stakeholders recognizing that OOS girls could not be characterized as a homogenous or uniform population.

### 3.2. Strategies for Enumerating OOS Girls

While stakeholders consistently described lack of consistent or systematic methods for enumerating OOS girls, they reported three main approaches for estimating the size of this population to guide HPV vaccination planning: national level population enumeration, household level enumeration, and proxy-based enumeration.

#### 3.2.1. National level population enumeration

Estimating the number of OOS girls is a crucial step for national-level HPV vaccination efforts, as it informs the quantity of vaccine needed for the program. Stakeholders commonly referenced national censuses to estimate the total population of target-aged girls. These data were often combined with school enrollment or attrition data collated by the Ministries of Education or derived from Demographic and Health Surveys (DHS) to infer the number of girls not in school, using simple calculations as below:

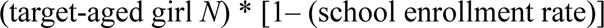

OOS girl N = *or*

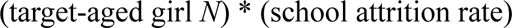

However, the quality of these data was widely perceived as questionable and inaccurate: “We use the census from 2019, but these numbers are estimates, and the quality isn’t great,” noted one Cambodian stakeholder. Likewise, in Uganda, the Ministry of Gender and Social Development maintained broader OOS population size estimates, but stakeholders still expressed concern over data accuracy and the lack of disaggregation by school-going status at the health facility level.

Others echoed that national statistics were not sufficiently granular to inform community-level planning, especially in rural or underserved areas, highlighting the need for more precise estimates of the OOS girl population. The lack of data transformability from national to the sub-national and community estimates was framed as a significant barrier to planning, monitoring, and evaluating HPV vaccine delivery strategies for OOS girls.

#### 3.2.2. Household level enumeration

With respect to subnational vaccination delivery planning, stakeholders stressed the value of direct enumeration through headcounts or name-based registries—particularly to increase accuracy of microplanning. In Kenya and Malawi, community health workers (CHWs), religious leaders, and volunteers visited households to identify OOS girls and recorded their names, ages, and school-going status. Community leaders’ knowledge regarding OOS girls and the reasons they did not attend schools played an essential role in individual-level identification. In Malawi, this information was compiled into separate lists of school-going and OOS girls, which were used to allocate local outreach resources. These lists were then distributed to schools and CHWs for follow-up vaccination.

> *“The identification of out-of-school girls… is dependent on these three [groups]: community health workers, volunteers, and the local leaders themselves. These are the ones that go around in the villages, especially the volunteers, to identify those out-of-school girls because they live within the same villages […] They know which girl doesn’t go to school and why.” –* Malawi

Although this approach was praised for its accuracy, it was considered labor- and resource-intensive. The consensus across countries was that enumeration efforts remain fragmented, under-resourced, and inconsistently applied. Many stakeholders expressed a desire for dedicated, standardized systems that could both enumerate and track OOS girls with greater precision—particularly in areas with large populations of zero-dose children (those who have not received a single dose of routine immunizations) or in areas with restricted access to healthcare. Kenya’s implementation of an electronic Community Health Information System (eCHIS) enabled CHWs to register girls digitally and monitor their vaccination status longitudinally. This system reportedly improved tracking and targeting during outreach. However, one Kenyan stakeholder noted that CHWs still “don’t know what our target population for out-of-school girls is,” as the digital system does not specify school-going status. Although it is useful for overall service delivery, eCHIS still requires accurate upfront identification of OOS girls prior to the implementation.

Stakeholders in Mozambique and Uganda emphasized the importance of leveraging local community knowledge to identify OOS girls, particularly in nomadic, refugee, or displaced communities. Uganda’s strategy involved empowering local leaders to work with CHWs to validate the lists of OOS girls and ensure comprehensiveness and inclusivity. Some stakeholders described efforts to reverify the existing manual headcounts annually or during sentinel health-seeking events like Child Health Days.

Despite these promising practices, headcount data was rarely digitized, standardized, or reported at a level beyond the individual facility or community. Consequently, the resource- and time-intensive preparation of these data were seldom leveraged for broader applications, and its utility beyond local planning varied across countries. Even when manual headcounts were conducted in communities, some stakeholders reported that the data were not aggregated beyond the community level, limiting their utility for planning or tracking vaccine coverage at regional or national scales. In contrast, one stakeholder in Malawi reported that community health teams map OOS girls by walking through villages with support from local leaders. They then aggregated this data at district and national levels to refine coverage estimates.

#### 3.2.3. Distinguishing between in-school and OOS girls in HPV vaccine delivery and evaluation

While in-school and OOS girls may be enumerated separately in the planning phase of HPV vaccination programs, the distinction becomes less clear during vaccine implementation and evaluation (see **Figure 2**). During delivery, many OOS girls in institutions, such as informal religious schools, orphanages, or correctional institutions, can be vaccinated through the institution-wide approaches that function similarly to school-based delivery. For instance, in Kenya, religious schools operating outside the Ministry of Education’s purview were registered to ensure their inclusion in school-based HPV vaccination delivery. Conversely, some in-school girls who missed school on vaccination day were vaccinated in health facilities or through community-based outreaches along with OOS girls. Therefore, delivery platforms often mix girls by schooling status, leading some stakeholders to question the operational value of this distinction. As one stakeholder explained,

> *“Those we vaccinate in school, we count as vaccinated in-school. Those in the community, they are being mixed up: those who are actually really out of school, and those who are in school but were not vaccinated in school.”* – Cameroon

**Figure 2.**
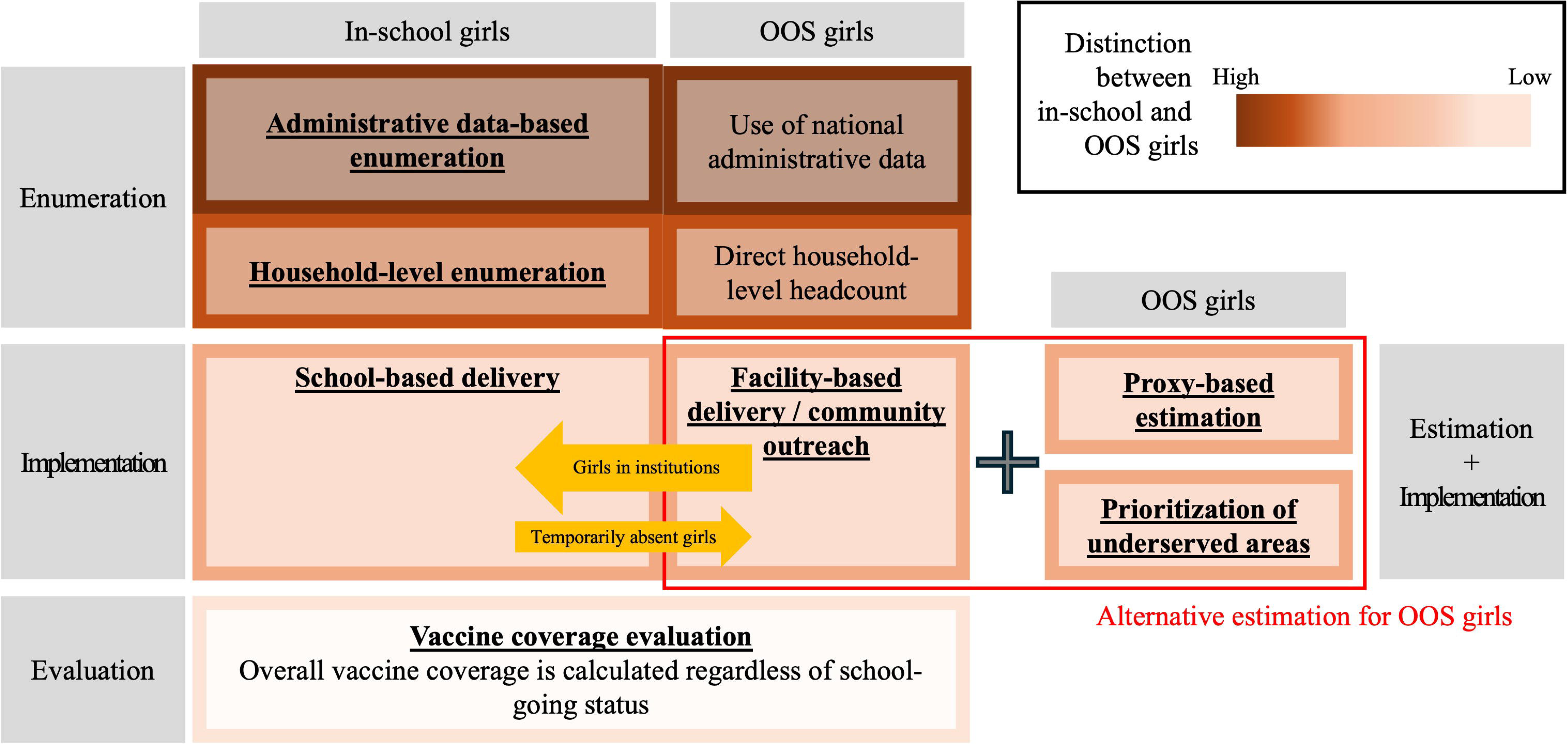
Enumeration methods of out-of-school (OOS) girls, and how these methods are used for the following implementation and evaluation phases

In the evaluation phase, stakeholders across countries similarly reported that school-going status has limited applicability. A stakeholder in Cameroon explained that the national vaccination record system does not distinguish between girls vaccinated in school and those vaccinated out of school, making it “complicated to know to what extent the program has affected out-of-school girls.” Echoing this concern, a stakeholder in Uganda noted that vaccination coverage is disaggregated by site of delivery rather than by school attendance. Together, these stakeholders emphasized that the lack of disaggregation by school-going status not only constrains accurate coverage estimation in OOS girls but also limits the ability to identify which subgroups of girls remain unreached. As another Ugandan stakeholder reported,

> *“Uganda has achieved the HPV1 (coverage for the first HPV dose) target for the region. But who is contributing to that coverage? Who are we leaving out? Is it a school-going child or an out-of-school child? I don’t have that answer.”* – Uganda

#### 3.2.4. Alternative methods for estimating and reaching OOS girls

Given the cost, feasibility, and operational utility of conventional enumeration methods, some countries pursued more pragmatic strategies to approximate the size of OOS girls. By relying on collective recall, informal lists, or proxy indicators, these countries sought to maintain some visibility of the OOS population without requiring full counts. In Malawi and Cameroon, outreach planners relied on the number of girls vaccinated in the community (i.e., not in school settings) in the prior year as a proxy for the OOS population. This approach assumes that girls vaccinated outside of school settings are likely to be OOS, though stakeholders acknowledged that this method can include in-school girls who missed vaccination at schools. Using historical vaccination data, these countries were able to at least generate rough community-level estimates of the OOS population size.

Several countries reported allocating additional resources for reaching OOS girls in underserved areas identified through equity mapping or zero-dose prevalence data, even though they did not estimate the actual population size. The underlying assumption was that OOS girls are more likely to reside in marginalized communities or harder-to-reach geographies. For example, stakeholders in Uganda described using equity mapping to pinpoint areas with low vaccination coverage, including for HPV vaccination—such as islands, mountainous regions, and pastoralist settlements. Similarly, countries often expected that areas with low vaccination coverage and high zero-dose prevalence were also remote and underserved, and therefore, likely to include more OOS girls.

> *“There is definitely some overlap between the geographies where we see higher zero-dose as well as expect more out-of-school, so we try to make sure that the numbers that we’re getting from the ground make sense in terms of the high-level data.”* — Cambodia

These approaches for approximating the number of girls needing vaccination do not directly account for school-going status.

## 4. Discussion

This qualitative study, based on interviews with key stakeholders from five African and one Asian countries, found that while all countries recognized the importance of reaching OOS girls to reduce vaccine inequities, their definition of OOS girls and strategies for enumerating them varied considerably across HPV vaccination programs. Although countries often relied on an administrative definition of OOS girls (i.e., those not enrolled in formal schools), in practice, the characteristics of OOS girls are more complex and context-dependent, varying across settings. National level enumeration was deemed essential, but all countries raised concerns about data quality and accuracy. Household level enumeration was also highlighted but framed financially and logistically impractical. To balance cost, feasibility, and operational utility in the implementation and evaluation phases of HPV vaccination programs, some countries reported employing alternative approaches to estimate OOS girls. These countries reported using proxy-based estimation relying on rough approximations or historical data or prioritizing underserved areas where they would assume a greater presence of OOS girls. For preparation of regional or district-level vaccine implementation, this approach was perceived as a good compromise between accuracy and resource intensity.

The quality of administrative data, such as population census estimates and school enrollment rates, needs to be improved to enable more accurate enumeration of age-eligible populations for HPV vaccination, including OOS girls. However, addressing these data gaps is a complex and long-term challenge, often constrained by limited resources and competing priorities. The adoption of Electronic Immunization Registries (EIRs)—such as digital registries and monitoring platforms—could provide a more practical pathway for improving accuracy in vaccine planning and delivery. EIRs are designed to record individualized health data [19], enabling providers and CHWs to register immunization records in a timely manner, thereby improving the availability of high-quality data [20]. Given they can register children as close to birth as possible and track them as a birth cohort, EIRs can also enhance immunization forecasting by providing accurate denominators and numerators for eligible populations in targeted areas [19,21]. EIRs would increase the efficiency of implementation planning by allowing stakeholders to identify whom to reach at various levels—village, regional, and national—regardless of school attendance status, and to accurately estimate coverage after implementation is completed [22]. Combined with sending SMS reminders, EIRs in Vietnam have improved childhood routine vaccine coverage [23]. In our study, stakeholders reported that eCHIS in Kenya improved data timeliness and the tracking of girls for HPV vaccination. To support HPV vaccine forecasting by school enrollment status, documenting whether a girl is in school or OOS in the system would help CHWs reach OOS girls, who may have fewer opportunities to be immunized against HPV relative to in-school girls. To enhance the use of EIR registration, some countries offer registration incentives [19]. For example, in Uruguay, EIR registration is linked to eligibility for school entry or other social programs [24]. Financial incentives are applied to EIR registration in Peru [25]. If this system can add information about school enrollment, which can be identified during the planning or implementation, it would help track OOS populations for both vaccination and school enrollment. Adoption of the EIR system and similar promising systems can improve national- and regional-level data accuracy.

Household level enumeration is an accurate method that can be used in settings where population data and registration is largely incomplete or outdated. Direct headcounts can complement microplanning by providing high-quality, up-to-date information on target populations. For instance, a study in Nigeria found that household-level enumeration identified 84% more individuals than census-derived estimates [26]. Direct household enumeration in hard-to-reach communities can also improve estimation and immunization coverage in these areas [27]. Because administrative data do not indicate individual school-going status, identifying OOS girls during community household enumeration enables accurate planning of HPV vaccination delivery in non-school settings. During the headcount process, typologies of OOS girls can be useful resources to help identify settings or venues where these girls congregate.

Ideally, HPV vaccination programs require accurate estimates of the target population—such as the number of eligible girls—to ensure sufficient vaccine supply and equitable coverage. However, in many resource-constrained settings, population enumeration for vaccine delivery poses substantial logistical and methodological challenges [7,28]. Furthermore, in contexts where no reliable population registry exists, and population mobility is high (e.g., pastoralist or nomadic communities), accurate estimation of vaccine-eligible girls is challenging [29]. As a result, our study found that some countries rely on low-granularity, proxy-based estimates to guide vaccine distribution. For example, the process of administering vaccines in the field and knowing how many doses were delivered in a given year can serve as a proxy for setting target doses in the following year, generating empirical information about the target population size. While the absence of accurate denominator data makes it difficult to assess true vaccination coverage or evaluate the equity of immunization efforts, this pragmatic approach supports rapid implementation and operational feasibility. The effectiveness of this approach, however, is predicated upon sufficient community mobilization and effective communication to ensure that eligible individuals are aware of—and seek out—HPV vaccination [30]. Furthermore, strategic selection of vaccination sites that maximize accessibility based on the typologies of OOS girls would be key to increase HPV vaccination among OOS girls with fewer resources. This approach allows vaccine delivery to function as both a service provision and a dynamic enumeration mechanism, effectively transforming an operational constraint into an opportunity to strengthen population data systems in harder-to-reach or rapidly evolving contexts.

Accurate enumeration of school enrollment status is critical during the planning phase of vaccination programs, as they directly inform the estimation of required doses and help to identify the target population for implementation. However, in many countries, these efforts do not extend consistently into the implementation and evaluation phases. During vaccine delivery in health facilities or community outreach, the target population often becomes less distinct, encompassing both OOS girls and those who missed vaccination opportunities in schools. Some girls in institutions (e.g., informal schools) may be vaccinated through school-based delivery efforts. As a result, any precision achieved during the planning stage becomes opaque in practice. Moreover, during evaluation, vaccination coverage is typically reported as an aggregate measure of total coverage, without disaggregating by school attendance status. This lack of differentiation limits the understanding of whether the program effectively reached the intended subgroups.

Disaggregating vaccination coverage by school attendance status can provide important insights into program reach and equity. If countries generate and report coverage estimates for OOS girls separately, it can enhance accountability and draw attention to this often under-reached population. Visibility of such data can also motivate vaccine implementers and local health authorities to identify barriers and strengthen strategies specifically aimed at improving HPV vaccination coverage among OOS girls. Accordingly, data disaggregation not only improves monitoring accuracy but also promotes more equitable implementation.

### 4.1. Recommendations

Balancing pragmatism with accuracy remains a central challenge in vaccine delivery in LMICs. To enhance HPV vaccination in the absence of supplemental resources, countries can leverage several strategies. Extending school-based delivery to informal schools or other institutions can improve coverage among girls who are often deemed OOS. Continued digitization of data platforms can streamline monitoring and reporting. Embedding HPV microplanning within broader microplanning for other antigens can help countries optimize limited resources and reduce costs, an especially important strategy given the significant resource constraints under the Gavi 6.0 framework [31]. Identifying and using typologies to prioritize areas with high concentrations of OOS girls can improve outreach efficiency.

Documenting school-going status on tally sheets—routine forms used to record administered doses—and carrying forward program records from previous years can support data-driven adjustments and prevent duplication in planning efforts.

### 4.2. Strengths and Limitations

This study offers a rare cross-country view of HPV vaccine implementation strategies for OOS girls, an understudied and hard-to-reach population. By incorporating perspectives from diverse national and sub-national stakeholders, it highlights a range of operational definitions, enumeration challenges, and community-level innovations in LMICs. These insights provide practical guidance for countries seeking to improve equity in HPV vaccine delivery. Nonetheless, our results are subject to some limitations. Findings are based on interviews with a limited number of stakeholders, and insights may not reflect all perspectives within countries or be transferable to countries not included in the present study. Stakeholder availability and language proficiency also limited representation to mostly African countries. Third, our ability to recruit participants from countries across different geographic regions and levels of program maturity was limited, primarily due to stakeholder availability, language barriers, and insufficient connections with local stakeholders. Lastly, despite our best efforts to assure participants of confidentiality and anonymity, some stakeholders may have self-censored and refrained from disclosing potentially sensitive information regarding outreach to OOS girls, especially in contexts where strategies were still pending or had not been successful. Furthermore, there may be inherent differences between stakeholders who chose to participate and those who were unavailable or unresponsive.

## 5. Conclusion

Reaching OOS girls is necessary to achieve the WHO target to vaccinate 90% of girls against HPV by the age 15, thereby preventing cervical cancer. Additionally, the enumeration and engagement of OOS girls is critical from an equity perspective, as they disproportionately come from families with fewer resources and are, therefore, rendered more vulnerable to adverse health outcomes [32]. Efforts to enumerate and reach OOS girls with HPV vaccination services can be especially resource-intensive, but they represent critical touchpoints for reducing future costs associated with cervical cancer diagnosis and treatment in unvaccinated or under-immunized populations. Understanding the characteristics of OOS girls and how programs practically identify them can help policymakers, healthcare providers, and program implementers tailor HPV vaccination programs to better reach OOS girls. Future studies are warranted to examine the practical utility and effectiveness of these alternative estimation methods in real-world settings, as well as to compare them with other enumeration strategies.

## Data Availability

All data produced in the present study are available upon reasonable request to the authors

## Acknowledgement

This work would not be possible without the national immunization stakeholders who shared their time and insights. We extend our gratitude to them. We also acknowledge collaborators from the HAPPI Consortium: JSI Research & Training Institute, Inc., Clinton Health Access Initiative, Jhpiego, PATH.

